# Importance of variables from different time frames for predicting self-harm using health system data

**DOI:** 10.1101/2024.04.29.24306260

**Authors:** Charles J. Wolock, Brian D. Williamson, Susan M. Shortreed, Gregory E. Simon, Karen J. Coleman, Rodney Yeargans, Brian K. Ahmedani, Yihe Daida, Frances L. Lynch, Rebecca C. Rossom, Rebecca A. Ziebell, Maricela Cruz, Robert D. Wellman, R. Yates Coley

**Author notes:** Corresponding author* : Charles J. Wolock, Department of Biostatistics, Epidemiology and Informatics University of Pennsylvania, 423 Guardian Drive, Philadelphia, PA, 19104, USA.

## Abstract

**Objective:** Self-harm risk prediction models developed using health system data (electronic health records and insurance claims information) often use patient information from up to several years prior to the index visit when the prediction is made. Measurements from some time periods may not be available for all patients. Using the framework of algorithm-agnostic variable importance, we study the predictive potential of variables corresponding to different time horizons prior to the index visit and demonstrate the application of variable importance techniques in the biomedical informatics setting.

**Materials and Methods:** We use variable importance to quantify the potential of recent (up to three months before the index visit) and distant (more than one year before the index visit) patient mental health information for predicting self-harm risk using data from seven health systems. We quantify importance as the decrease in predictiveness when the variable set of interest is excluded from the prediction task. We define predictiveness using discriminative metrics: area under the receiver operating characteristic curve (AUC), sensitivity, and positive predictive value.

**Results:** Mental health predictors corresponding to the three months prior to the index visit show strong signal of importance; in one setting, excluding these variables decreased AUC from 0.85 to 0.77. Predictors corresponding to more distant information were less important.

**Discussion:** Predictors from the months immediately preceding the index visit are highly important. Implementation of self-harm prediction models may be challenging in settings where recent data are not completely available (e.g., due to lags in insurance claims processing) at the time a prediction is made.

**Conclusion:** Clinically derived variables from different time frames exhibit varying levels of importance for predicting self-harm. Variable importance analyses can inform whether and how to implement risk prediction models into clinical practice given real-world data limitations. These analyses be applied more broadly in biomedical informatics research to provide insight into general clinical risk prediction tasks.

## 1. INTRODUCTION

Preventing fatal and non-fatal self-harm is a public health priority: In the United States alone, in 2021 over 48,000 people died by suicide and an estimated 1.7 million adults attempted suicide [1, 2]. Health care settings provide an opportunity to prevent self-harm behavior if those at higher risk can be accurately identified. Health system data, including electronic health records (EHR) and health insurance claims data, contain detailed clinical history relevant to mental health risk factors and other predictors of self-harm. Additionally, identification of self-harm risk using health system data enables implementation of risk prediction models within EHR platforms for clinical use. Several models have been developed using health system data to predict the risk of fatal and non-fatal self-harm [3–14]. Many of these prediction models achieve an area under the receiver operating characteristic curve (AUC) of 0.8 or above, implying good prediction performance as measured by discrimination between those who do and those who do not attempt or die by suicide. Self-harm risk varies over time; suicidal ideation, depressive symptoms, and other factors are not static. Many clinical prediction models for self-harm risk use information available prior to a medical visit to assess a patient’s risk. We refer to the visit at which the prediction is made as the *index visit*. For predictors that can vary over time, it is common for these models to use information from up to five years prior to an index visit [7, 14–17]. In most cases, the predictors are divided into several overlapping time intervals: for example, predictors corresponding to information from the 90 days, one year, and five years prior to the visit [14, 15, 17], or 30 days, 90 days, and one year prior to the visit [6].

Information measured close in time to a given visit, including recent diagnoses and dispensed prescriptions, is likely correlated with an individual’s current risk. However, it can be difficult to incorporate recent information into prediction models in real time. For example, there are often time lags in processing pharmacy or insurance claims data, especially from external providers, which means these data might not be available for risk prediction at the time of the index visit. Data from the Centers for Medicare and Medicaid Services on the time elapsed between delivery of health care service and claim submission indicate significant delays in submission of fee-for-service claims; only at three months after the date of service do submission rates for all claim types exceed 90% [18]. In health system–based prediction models, predictors are often defined and coded as the presence of an event (e.g., an inpatient mental health encounter during the past three months). A value of zero could indicate either a true absence (i.e., an individual has truly not had an inpatient mental health encounter in the past three months) or could reflect that the relevant data from that encounter has not yet been processed. This may be the case, for example, in a health system without inpatient facilities. Because of this lag, the health record for such a patient may contain inaccurate information at the time of the index visit; this may be a concern when deploying a risk prediction model whose performance was evaluated using retrospective cohort data. An additional concern is that some people have shorter clinical history available for prediction because they are new to the health system; health care information prior to enrollment in the current health system may be invisible to a prediction model. Shorter duration of clinical history could reflect less access to continuous insurance coverage and could be related to social determinants of health. Thus, it is of interest to determine the predictiveness of variables from particular time frames.

The statistical framework of *variable importance* can be used to investigate the predictiveness of a variable or group of variables. Variable importance can be broadly classified as either specific or agnostic to the algorithm used to construct the prediction model. Algorithm-specific variable importance measures (VIMs) quantify how the particular fitted algorithm uses variables to make predictions. Examples include the Gini criteria VIM returned by random forests algorithms [19], coefficients in penalized regression models [20], and changes in the prediction output by a model when certain variables are treated as missing [see, e.g., 21]. Algorithm-agnostic variable importance, in contrast, is the change in population prediction performance when certain variables are excluded from the model [see, e.g., 22–24]. Because algorithm-agnostic importance is not tied to a particular modeling strategy, its interpretation does not depend on the prediction technique used. Furthermore, by treating variable importance as a population quantity, the algorithm-agnostic approach allows for statistical inference. Both types of variable importance can provide complementary information [25]. However, the chosen VIM should reflect the scientific question at hand.

Our goal in this work is to understand the implications of using different subsets of temporally defined variables to develop prediction models for fatal and non-fatal self-harm using health system data. In particular, we aim to quantify improvement in self-harm prediction attributed to the inclusion of clinically derived variables from more recent (0–3 months) and distant (13–60 months) time periods preceding the index visit. We judged the algorithm-agnostic variable importance approach to be most appropriate for this task, because our goal is to understand the implications of using different subsets of variables to develop prediction models, rather than to understand how any given prediction model makes use of the variables it is provided. The analytic approach to variable importance taken here can be applied more broadly in biomedical informatics research to provide insight into clinical risk prediction tasks.

## 2. MATERIALS AND METHODS

### 2.1. Setting and study sample

Data for this study were collected from seven integrated health systems (HealthPartners, Henry Ford Health, and the Colorado, Hawaii, Northwest, Southern California, and Washington regions of Kaiser Permanente) that provide insurance coverage and comprehensive medical care to defined patient populations. Health system data, including EHR and insurance claims data, were extracted via each site’s research data warehouse [26]. Responsible Institutional Review Boards for each health system granted waivers of consent to use de-identified records data for this research.

The study sample included two categories of visits made by members aged 11 or older: mental health specialty visits (mental health setting) and general medical visits in which a mental health diagnosis was recorded (general medical setting). There were no eligibility requirements related to prior health insurance plan enrollment or health care utilization. All eligible visits from January 1, 2009 to September 30, 2017 were included in the study sample with the exception of visits from Henry Ford Health, which only contributed data following the implementation of a new electronic records system on January 1, 2012. Members may have had multiple eligible visits in either the mental health or general medical setting during this time period and, as such, multiple visits per member were included in the analytic sample.

### 2.2. Outcomes and follow-up

We separately considered prediction of two binary outcomes: any self-harm (including fatal and non-fatal) and suicide death (i.e., fatal self-harm only) within 90 days of an eligible visit. As in prior work, non-fatal self-harm events were ascertained from health system data, either the EHR or claims data, by identifying all ICD-9/10 injury or poisoning diagnoses accompanied by a cause of injury code indicating intentional self-harm or undetermined intent [8]. Suicide deaths were ascertained from state mortality records and were identified by a cause-of-death code for self-inflicted injury or injury or poisoning with undetermined intent [27, 28]. Diagnosis code lists are available online at https://github.com/MHResearchNetwork/more-srpm/blob/main/SRS3_DX_CODES_20181204.sas.

Prediction models for any self-harm excluded patients who were not enrolled in the health system’s insurance plan on the index date or for the following 90 days to enable complete outcome capture from insurance claims data. Insurance plan enrollment was not required for inclusion in suicide death prediction models, as mortality records data were available on all patients regardless of their current enrollment status. The study sample for suicide death prediction excluded visits that occurred after availability of cause-of-death data at each site (Supplementary Table S1).

### 2.3. Self-harm risk predictors

Predictors of self-harm were extracted from EHR and insurance billing information. ‘Base’ predictors included in all prediction models, but not assessed for variable importance, included age, sex, race, ethnicity, insurance type, and census-derived sociodemographic variables.

All other predictors were related to mental health diagnoses or mental health care utilization. Mental health–specific predictors covering the 0–3 months, 4–12 months, and 13–60 months prior to the visit included binary indicators of the following in each time period: mental health and substance use diagnoses (e.g., depression, anxiety, alcohol use disorder); dispensed psychiatric medications (e.g., antidepressants, benzodiazepines); prior outpatient, inpatient, and emergency department encounters with mental health diagnoses; prior suicide attempt and self-harm diagnoses; and responses to the Patient Health Questionnaire 9^th^ item [3, 15, 29, 30], which asks about suicidal ideation. A complete list of predictors is given in Supplementary Table S2.

### 2.4. Statistical analysis

We assessed variable importance by computing the difference in predictiveness (see predictiveness measures below) between pairs of fitted prediction models. The models considered are depicted in Figure 1. For a given predictor group of interest, the difference in predictiveness between a *larger model* (which includes the predictor group of interest and others) and *reduced model* (which excludes the predictor group of interest) quantifies how much predictiveness is lost by excluding predictors measured in a particular time period. This decrease in predictiveness is the variable importance of the predictor group relative to the full set of predictors in the larger model. Each model included the base predictors and some combination of month 0–3 predictors, month 4–12 predictors, and month 13–60 predictors. The models compared were:

– a model using months 0–12 was compared to a model using months 4–12 to assess the importance of month 0–3 predictors relative to month 0–12 predictors;
– a model using months 0–60 was compared to a model using months 4–60 to assess the importance of month 0–3 predictors relative to month 0–60 predictors;
– a model using months 4–60 was compared to a model using months 4–12 to assess the importance of month 13–60 predictors relative to month 4–60 predictors; and
– a model using months 0–60 was compared to a model using months 0–12 to assess the importance of month 13–60 predictors relative to month 0–60 predictors.

**Figure 1:**
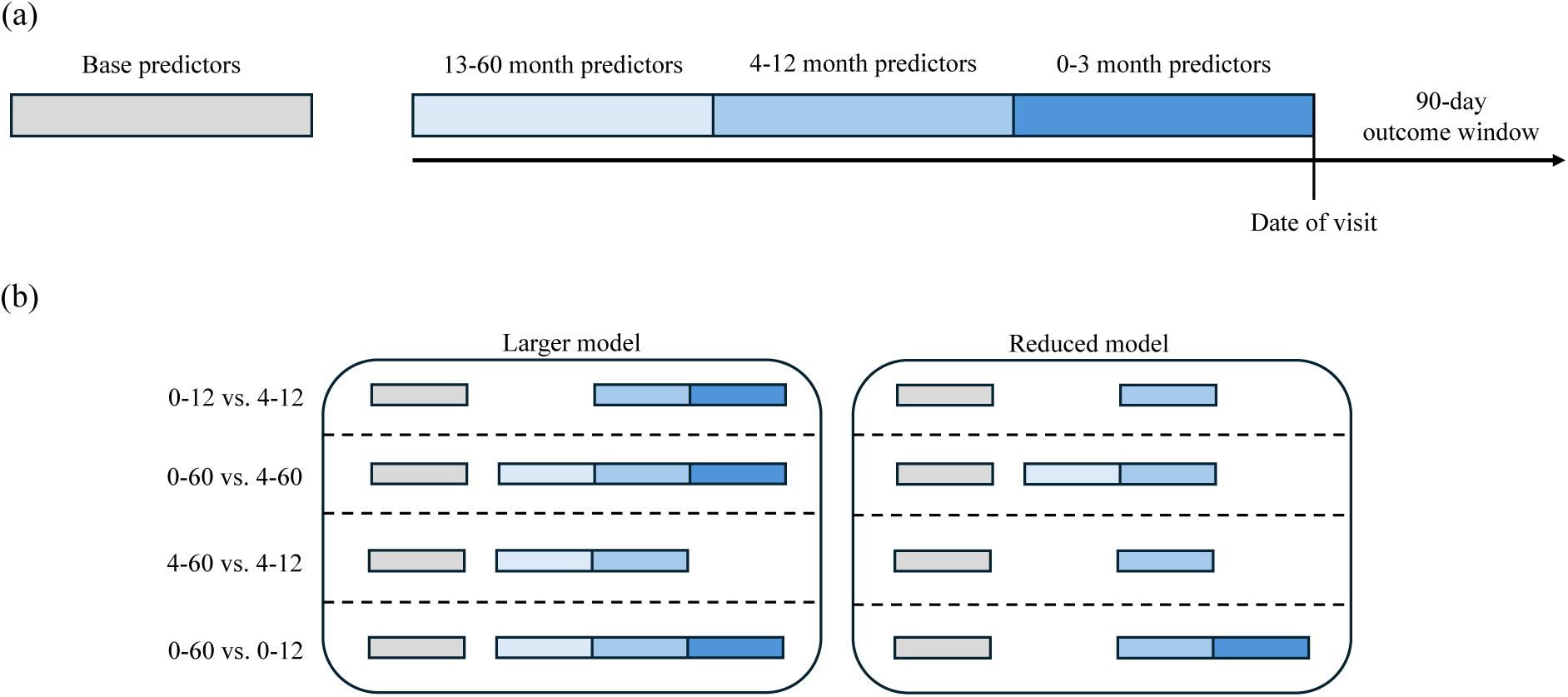
Schematic of temporal predictor groups in the variable importance analysis. (a) Predictors were categorized into four groups: base predictors, including demographics and comorbidities, that were included in all prediction models (gray), and mental health–specific predictors covering the 0–3 months (dark blue), 4–12 months (medium blue), and 13–60 months (light blue) prior to the prediction instance (vertical black line). The outcome window spanned 90 days from the prediction instance (date of the visit). Note that the timeline is not drawn to scale. (b) We made four comparisons to assess variable importance. In each case, the larger model used base predictors plus some subset of temporal predictors. The reduced model was constructed by removing a temporal predictor group.

Variable importance was quantified using predictiveness measures corresponding to AUC [31], sensitivity, and positive predictive value (PPV). Sensitivity and PPV were calculated using cutpoints based on the 90^th^, 95^th^, and 99^th^ percentiles of predicted risk. These quantities measure discriminative performance. This choice of predictiveness measures accords with the practical usage of the models to identify a subset of patients at higher risk; the methods described here could be applied using other measures.

We assessed variable importance in four setting-outcome pairs defined by visit type (mental health specialty vs. general medical) and outcome (any self-harm event vs. suicide death). For each, a sample of patient-visits was constructed according to visit type and outcome-specific inclusion criteria (described above). For each of the four outcome-setting pairs, we performed the procedure described below. Our approach leverages a cross-fitting procedure with validation of model performance in independent testing sets. This combination of techniques reduces bias due to potential overfitting of prediction models and enables robust inference on variable importance [24]. The procedure was as follows:

1. The full collection of patients contributing visits to each sample was randomly subdivided on the person level into five folds, which we refer to as *cross-fitting* folds. Cross-fitting, which entails training the prediction model and evaluating variable importance on separate subsets of data, has been shown to improve performance in variable importance analyses [24, 32].
2. Three cross-fitting folds were designated as training data; the remaining two were designated as test data.
3. Visits corresponding to patients in the training data were used to construct penalized logistic regression models via the lasso [33]. The lasso is a regression method that combines shrinkage of coefficients towards zero and exclusion of variables with estimated null coefficients from the prediction model. The lasso penalization parameter was selected via 10-fold cross-validation [34] within the training data (the set of three cross-fitting folds from step 2) using AUC loss, with cross-validation folds defined on the person level (rather than the visit level) to ensure independence between folds.
4. The fitted lasso model was used to generate cross-fit predicted probabilities for the visits corresponding to patients in the test data (the set of two cross-fitting folds from step 2).
5. Sample-split predictiveness estimates of the two models being compared were computed separately on the two independent folds comprising the test sample. This permits valid inference even under zero importance, as evaluating both models in the same sample risks type I error inflation [24, 35]. VIM estimates, given by the difference in estimated predictiveness of the two models, were truncated at zero. (The VIM parameter takes non-negative values by construction; increasing the number of available predictors cannot reduce model performance on a population level.) Variance estimates for predictiveness were computed separately on the two test folds using the nonparametric bootstrap [36] with 500 bootstrap replicates, resampled at the patient level. The variance of the VIM estimator was computed as the sum of variance estimates constructed from the two independent test folds [24].
6. To increase robustness against the random splitting of the data into folds, Steps 2–5 were repeated ten times, with different combinations of cross-fitting folds designed as training and test data. VIM estimates and corresponding variance estimates were averaged over all ten test/train combinations to give the final results. The final variance estimates were used to construct 95% confidence intervals based on a normal approximation.

The penalized logistic regression modeling approach was chosen due its strong performance in prior evaluations of self-harm risk prediction models in a similar patient population; more complex models such as random forests and artificial neural nets were found to have similar predictive performance [14].

In addition to the four primary setting-outcome pairs, we also performed analyses stratified by race and ethnicity. For these subgroup analyses, we considered only self-harm events and not suicide deaths due to the small number of suicide deaths in some subgroups. The penalized logistic regression prediction model was as described above, while the predictiveness estimates were computed only among visits corresponding to individuals reporting the race/ethnicity subgroup under consideration. We used AUC and sensitivity at the 90^th^, 95^th^, and 99^th^ race/ethnicity-specific percentiles of predicted risk. Because PPV varies depending on the event rate, comparisons of PPV across racial and ethnic subgroups are not informative [17], and we did not consider importance quantified in terms of PPV.

## 3. RESULTS

A total of 15,986,946 mental health visits made by 1,590,002 patients and 11,104,580 general medical visits made by 2,732,786 patients were included in our analysis. The rates of self-harm and suicide death in the 90 days following a visit were 0.64% and 0.023%, respectively, in the mental health sample and 0.33% and 0.016% in the general medical sample. Overall characteristics of the study sample are summarized in Table 1, and a subset of the temporal predictors are summarized in Table 2.

**Table 1:** Characteristics of the patient visits included in the study, summarized by visit type (mental health specialty or general medical). ^a^Patients may have multiple types of insurance. ^b^Percentage calculated using only visits included in the analysis.

|  | Mental health<br>*n* = 15,986,946 | General medical<br>*n* = 11,104,580 |
| --- | --- | --- |
|  | *n* (%) | *n* (%) |
| Female | 10,173,202 (63.6%) | 6,964,601 (62.7%) |
| Age, years |  |  |
| 11–17 | 1,762,956 (11.0%) | 692,306 (6.2%) |
| 18–29 | 2,700,008 (16.9%) | 1,418,491 (12.8%) |
| 30–44 | 4,068,753 (25.5%) | 2,191,953 (19.7%) |
| 45–64 | 5,601,992 (35.0%) | 3,832,204 (34.5%) |
| 65 and older | 1,853,237 (11.6%) | 2,969,626 (26.7%) |
| Race, self-reported |  |  |
| American Indian/Alaskan Native | 152,863 (0.96%) | 125,382 (1.1%) |
| Asian | 785,358 (4.9%) | 522,750 (4.7%) |
| Black/African American | 1,393,712 (8.7%) | 868,921 (7.8%) |
| Native Hawaiian/Pacific Islander | 169,010 (1.1%) | 101,295 (0.91%) |
| White | 10,858,962 (67.9%) | 7,808,508 (70.3%) |
| Multiple or other races indicated | 85,075 (0.53%) | 95,466 (0.86%) |
| Hispanic/Latino ethnicity | 3,876,798 (24.2%) | 2,384,030 (21.5%) |
| No race or ethnicity recorded | 615,203 (3.8%) | 384,300 (3.5%) |
| Insurance<sup>a</sup> |  |  |
| Commercial group | 11,669,276 (73.0%) | 6,698,458 (60.3%) |
| Individual | 2,260,530 (14.1%) | 2,017,579 (18.2%) |
| Medicaid | 1,083,167 (6.8%) | 978,177 (8.8%) |
| Medicare | 2,567,666 (16.1%) | 3,313,068 (29.8%) |
| Any fatal or non-fatal self-harm |  |  |
| # visits included in analysis | 15,249,031 (95.4%) | 10,551,857 (95.0%) |
| # visits with 90-day event<sup>b</sup> | 98,089 (0.64%) | 34,764 (0.33%) |
| Suicide death |  |  |
| # visits included in analysis | 13,981,418 (87.5%) | 9,714,817 (87.5%) |
| # visits with 90-day event<sup>b</sup> | 3,199 (0.023%) | 1,510 (0.016%) |

**Table 2:** Summary of selected temporal predictors for patient visits included in the study. MH: mental health.

|  | Mental health<br>*n* = 15,986,946 | General medical<br>*n* = 11,104,580 |
| --- | --- | --- |
|  | *n* (%) | *n* (%) |
| Depression diagnosis |  |  |
| 0–3 months | 8,515,731 (53.3%) | 2,757,582 (24.8%) |
| 4–12 | 7,443,083 (46.6%) | 3,815,869 (34.4%) |
| 13–60 months | 7,884,537 (49.3%) | 4,964,064 (44.7%) |
| Anxiety diagnosis |  |  |
| 0–3 months | 8,041,059 (50.3%) | 2,525,512 (22.7%) |
| 4–12 months | 7,110,947 (44.5%) | 3,242,979 (29.2%) |
| 13–60 months | 7,555,451 (47.3%) | 4,478,364 (40.3%) |
| Antidepressant fill |  |  |
| 0–3 months | 7,967,051 (49.8%) | 4,013,619 (36.1%) |
| 4–12 months | 7,731,060 (48.4%) | 4,531,589 (40.8%) |
| 13–60 months | 7,993,183 (50.0%) | 5,333,794 (48.0%) |
| Benzodiazepine fill |  |  |
| 0–3 months | 3,888,653 (24.3%) | 1,969,251 (17.7%) |
| 4–12 months | 4,229,825 (26.5%) | 2,393,763 (21.6%) |
| 13–60 months | 5,421,777 (33.9%) | 3,552,294 (32.0%) |
| Inpatient MH encounter |  |  |
| 0–3 months | 1,112,531 (7.0%) | 587,133 (5.3%) |
| 4–12 months | 1,309,140 (8.2%) | 762,549 (6.9%) |
| 13–60 months | 2,377,345 (14.9%) | 1,629,003 (14.7%) |
| Emergency department MH encounter |  |  |
| 0–3 months | 1,746,647 (10.9%) | 1,004,143 (9.0%) |
| 4–12 months | 2,099,814 (13.1%) | 1,255,128 (11.3%) |
| 13–60 months | 3,548,115 (22.2%) | 2,337,818 (21.1%) |
| Prior self-harm |  |  |
| 0–3 months | 199,478 (1.2%) | 41,295 (0.37%) |
| 4–12 months | 201,901 (1.3%) | 55,047 (0.50%) |
| 13–60 months | 348,536 (2.2%) | 130,139 (1.2%) |
| PHQ 9th item response 2 or 3 |  |  |
| 0–3 months | 546,085 (3.4%) | 81,294 (0.73%) |
| 4–12 months | 489,087 (3.1%) | 110,727 (1.0%) |
| 13–60 months | 398,428 (2.2%) | 147,768 (1.3%) |

Performance of risk prediction models using predictors from all time periods is reported in Table 3. The AUC estimates range from 0.807 to 0.850, with superior performance observed for predicting any self-harm versus suicide death. We observe a similar pattern for sensitivity. For example, sensitivity using the 95th risk score percentile cut-point was 45.7% for predicting any selfharm following a mental health specialty visit and 47.2% for predicting any self-harm following a general medical visit; for predicting suicide death, these values were 35.1% and 38.4%, respectively. PPV for predicting suicide death was low, in keeping with the low prevalence of fatal self-harm. Overall, these estimates are similar to those observed in previous studies of self-harm risk prediction in this setting [8], suggesting that the fitted lasso models perform as expected.

**Table 3:** Performance of any self-harm (fatal and non-fatal) and suicide death prediction models including predictors from all time periods.

|  | Mental health visits |  | General medical visits |  |
| --- | --- | --- | --- | --- |
|  | Any self-harm | Suicide death | Any self-harm | Suicide death |
| AUC | 0.850 | 0.807 | 0.829 | 0.815 |
| Sensitivity (%) |  |  |  |  |
| 90th percentile | 60.2 | 50.0 | 59.4 | 51.0 |
| 95th percentile | 45.7 | 35.1 | 47.2 | 38.4 |
| 99th percentile | 19.0 | 10.9 | 23.5 | 18.9 |
| PPV (%) |  |  |  |  |
| 90th percentile | 3.8 | 0.10 | 1.9 | 0.08 |
| 95th percentile | 5.7 | 0.14 | 3.0 | 0.11 |
| 99th percentile | 11.6 | 0.21 | 7.6 | 0.28 |

Figures 2–4 show the variable importance results for predictiveness measures corresponding to AUC, sensitivity, and PPV, with the latter two measures evaluated using the 95th risk score percentile cut-point. Additional results for sensitivity and PPV at other cut-points are given in the Supplementary Material.

**Figure 2:**
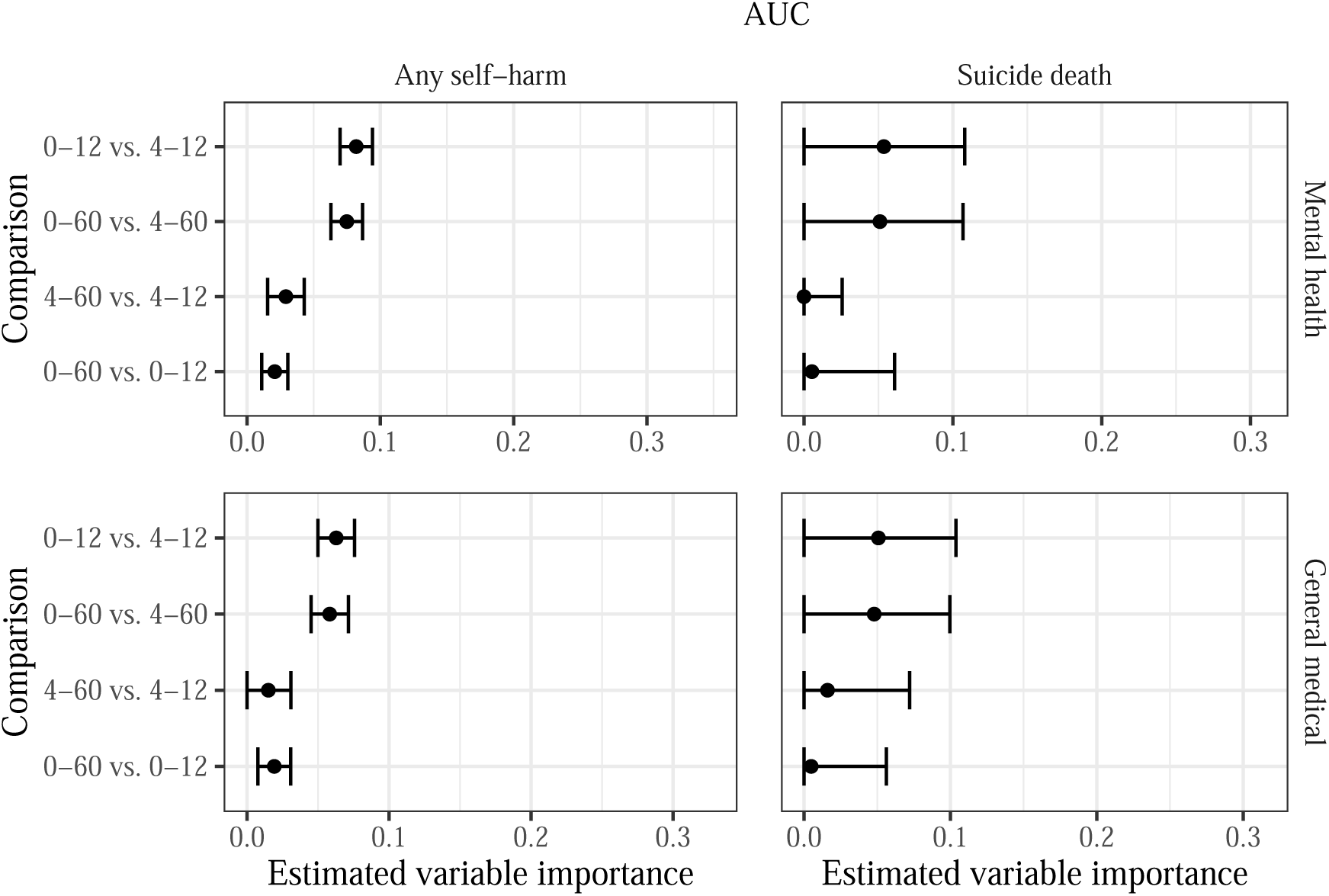
Estimated variable importance for temporal predictor groups in terms of **AUC**. Note the different *x*-axis scales for each outcome-setting pair, which are based on the estimated maximum possible variable importance (see Supplementary Material for details).

**Figure 3:**
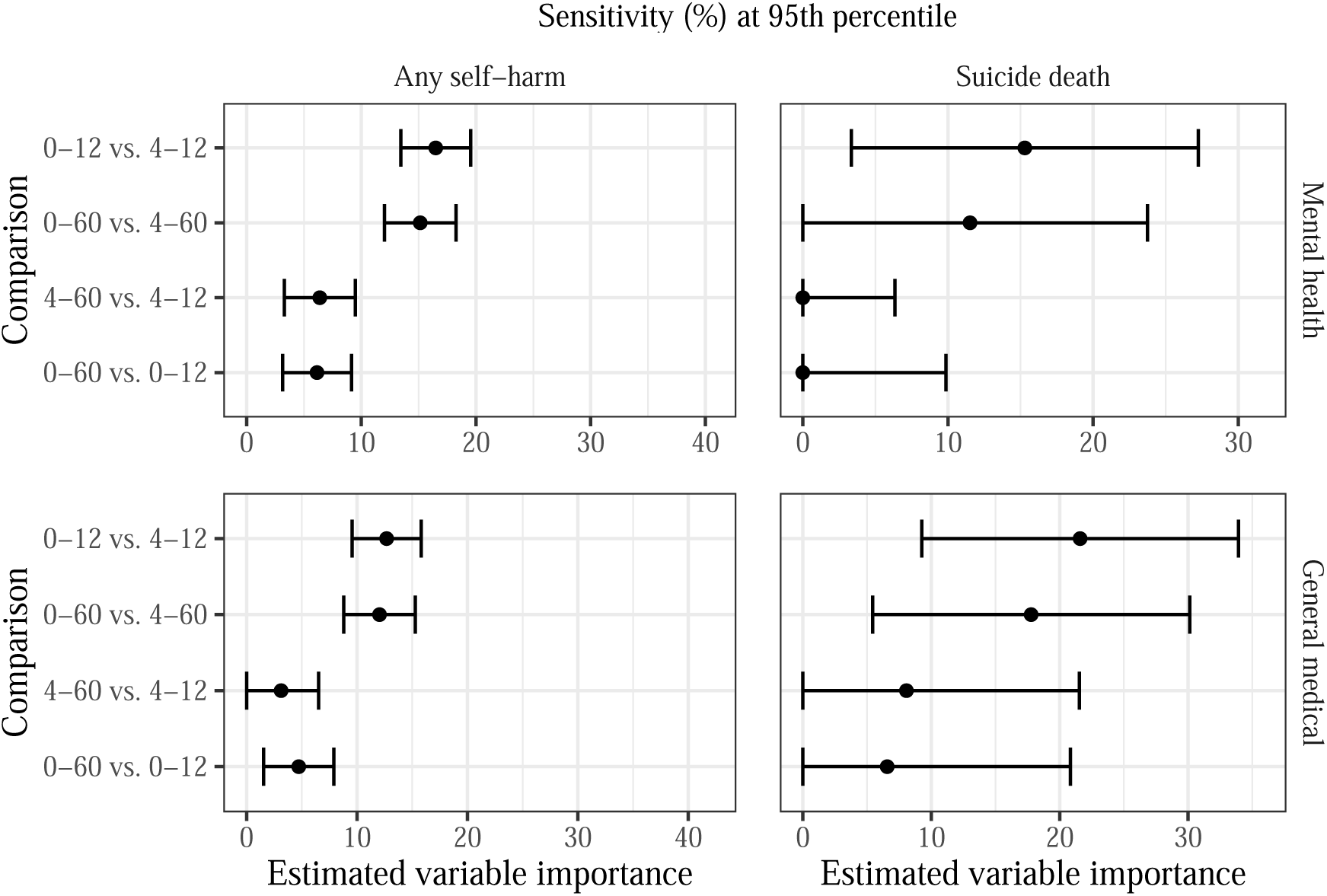
Estimated variable importance for temporal predictor groups in terms of **sensitivity at the 95th percentile of risk scores**. Note the different *x*-axis scales for each outcome-setting pair, which are based on the estimated maximum possible variable importance (see Supplementary Material for details).

**Figure 4:**
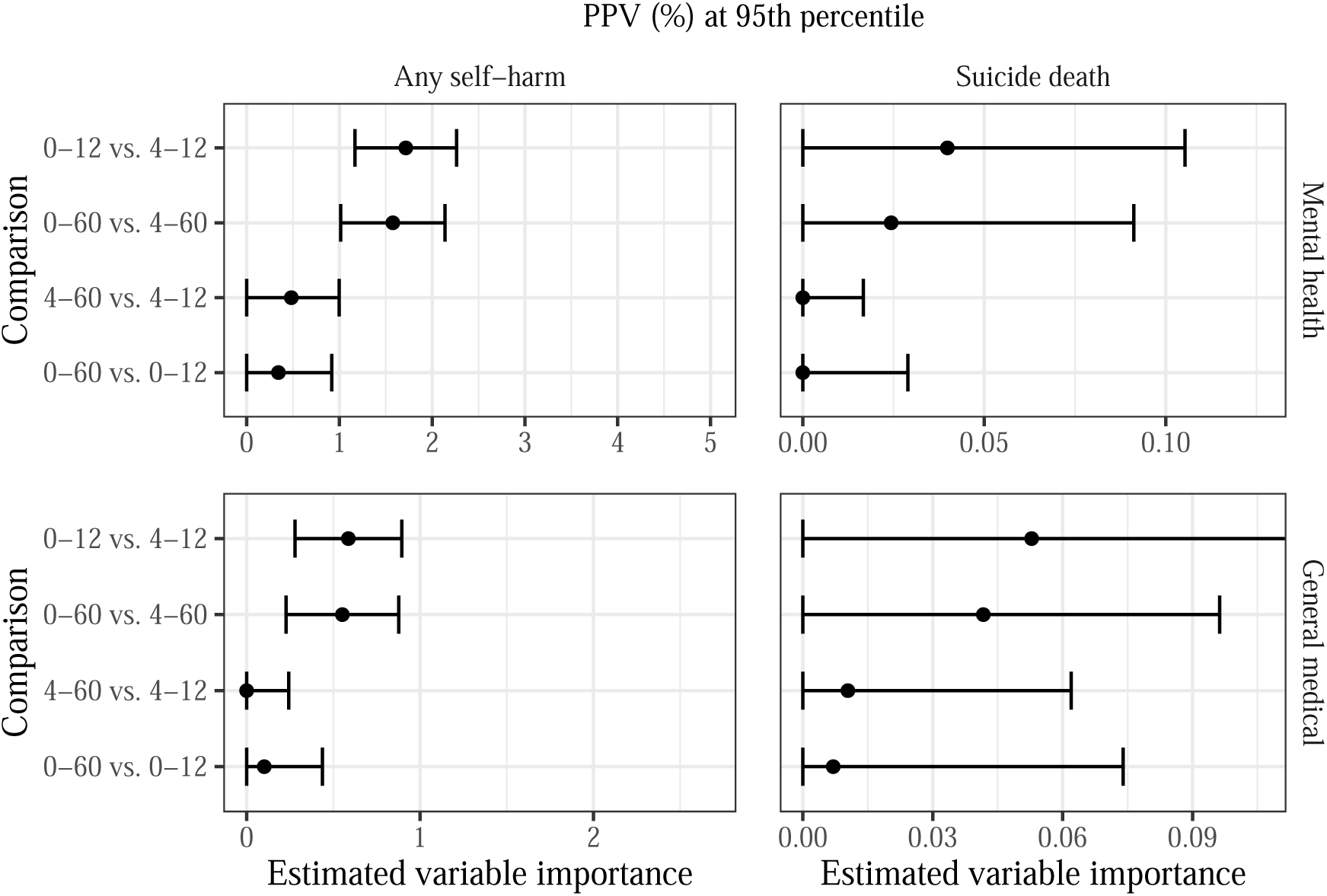
Estimated variable importance for temporal predictor groups in terms of **PPV at the 95th percentile** of risk scores. Note the different *x*-axis scales for each outcome-setting pair, which are based on the estimated maximum possible variable importance (see Supplementary Material for details).

In Figure 2, we show the variable importance results for AUC predictiveness. Focusing first on the top left panel of Figure 2, we observe that the most recent predictors, capturing information from 0–3 months prior to the visit, show strong signal of importance for predicting any self-harm following a mental health visit. Compared to a model using predictors from months 0–60, the model using only months 4–60 shows substantially worse risk discrimination, with a decrease in AUC from 0.850 to 0.776 (VIM = 0.075, 95% CI 0.063–0.087). Likewise, removing the month 0–3 predictors from a model using months 0–12 results in a drop in AUC from 0.846 to 0.764 (VIM = 0.082, 95% CI 0.070–0.094). Predictors from 13–60 months appear somewhat less important: Comparing the 0–60 month model to the 0–12 month model corresponds to a decrease in AUC from 0.850 to 0.830 (VIM = 0.021, 95% CI 0.011–0.031), and comparing the 4–60 month model to 4–12 months corresponds to a decrease in AUC from 0.793 to 0.764 (VIM = 0.029, 95% CI 0.015–0.043).

We observe similar patterns for sensitivity (top left panel of Figure 3) and PPV (top left panel of Figure 4) using the 95th risk score percentile cut-point. Removing months 0–3 from a larger prediction model results in a drop in sensitivity from 45% to between 28% and 30%, corresponding to a VIM value between 15% and 17%. Removing months 13–60, conversely, decreases sensitivity by only 6%. There is strong evidence of non-zero importance for the variable groups in question for each of these model comparisons. For PPV, we observe a drop from between 5.6% and 5.7% to between 3.8% and 4.0% when removing predictors from months 0–3, while the decrease from removal of months 13–60 is only 0.4%.

The bottom left panels of Figures 2–4 show the results for predicting the risk of any self-harm after a general medical visit. By and large, the results mirror those seen in the mental health setting, particularly for AUC and sensitivity. For PPV, the magnitude of estimated importance is lower for all model comparisons and is near zero for the month 13–60 variables. The smaller magnitude of estimated importance matches the lower event rate for any self-harm after a general medical visit versus a mental health specialty visit.

The right columns of Figures 2–4 show the estimated variable importance for predicting the risk of suicide death. Compared to the inferential results for predicting any self-harm, there is substantially more uncertainty in the suicide death analyses due to the smaller number of fatal self-harm events observed in the data set. The overall pattern of variable importance is similar between mental health and general medical visits. For AUC variable importance, the estimated importance of months 0–3 is around 0.05, corresponding to a decrease in AUC from 0.81 to 0.76, and the estimated importance of months 13–60 is close to zero. For sensitivity, predictors from months 0–3 have fairly large estimated importance relative to months 0–12 in both mental health (VIM = 15.3%, 95% CI 3.3%–27.2%) and general medical (VIM = 21.6%, 95% CI 9.3%–33.9%) settings. Month 0–3 predictors also demonstrate substantial importance relative to months 0–60 in the general medical setting (VIM = 17.8%, 95% CI 5.4%–30.1%). The ranking of variable importance estimates in terms of PPV is similar for predicting suicide death as for predicting any self-harm, although the confidence intervals are wide due to the low event rate and small absolute number of events.

In the Supplementary Material, we present results for sensitivity and PPV at cut-points based on the 90th and 99th percentiles of estimated risk; the overall patterns mimic those seen in Figures 3 and 4. The magnitude of the estimated VIMs varies by the percentile of risk score used as a cut-point. For sensitivity, for example, using a lower cut-point results in both higher sensitivity for all models (see Table 3) and larger VIM values, i.e., larger absolute differences between models. The opposite pattern is observed for PPV variable importance because PPV decreases as more visits are classified as high-risk.

The Supplementary Material also contains results for variable importance analyses stratified by self-reported race and ethnicity. Overall model performance using predictors from all time periods is similar across subgroups. Compared to the unstratified analyses, the variable importance results are broadly similar, with VIMs for months 0–3 estimated to be larger in magnitude than VIMs for months 13–60. However, there is substantially higher estimated variability in the subgroup analyses, as evidenced by wider confidence intervals.

## 4. DISCUSSION

Using a sample of over 27 million visits made by patients across seven health systems, we assessed the importance of temporally grouped sets of variables for predicting the 90-day risk of any self-harm (fatal and non-fatal) and of suicide death. Using several predictiveness measures, we found consistent evidence that mental health–specific features corresponding to the most recent three months prior to the visit were highly important for predicting the risk of any self-harm. We found slightly weaker evidence for the importance of these features in predicting the risk of suicide death, although there was substantially larger uncertainty in evaluating the suicide death prediction models due to the smaller number of events. For prediction of any self-harm following a mental health specialty visit, removing predictors from the most recent three months resulted in a drop in AUC from 0.85 to 0.78, corresponding to a loss of nearly 20% of the discriminative potential of the model relative to the AUC of a null model (0.5). Features capturing patient information from one to five years prior to the prediction instance appear less important than more recent features.

One concern motivating this study was that complete information on recent predictors may not be available in real-time to include in risk calculations. This may be due to delays in processing health insurance claims: Information that may only be available in claims data (such as prescription fills or encounters with providers external to the health system) will not be immediately reflected in the health system data used to generate risk predictions at the time of the index visit. The impact of this lag cannot be easily examined in prediction modeling studies with retrospective cohort data, since it is challenging to determine what data would have been available at the time of an encounter. Thus, we used variable importance analyses to examine the predictive contribution of risk factors in the three months preceding a visit. This three month window reflects a time period after which most claims data are expected to be available; for instance, over 90% of fee-for-service Medicare and Medicaid claims are submitted within three months [18]. We found these recent predictors to be highly important, indicating that a model excluding this information would not as accurately identify patients at higher risk of self-harm following a visit. This result suggests that realizing good real-time predictive performance of self-harm prediction models would be unlikely for claims-only settings where recent data are not completely available at the time a prediction is made. In a health system with access to clinical and claims data, we recommend prospectively monitoring availability of risk factors and performance of models with real-time data to quantify the impact of delayed data availability on identification of high-risk visits.

A second concern motivating this study was that patients without long-term, sustained insurance coverage would not have complete information on self-harm risk factors going back five years. Stable insurance coverage not only influences access to affordable health care but is also related to other social determinants of health including employment security and financial strain. As such, implementing a self-harm prediction model with differential capture of predictors could exacerbate existing health disparities, a concern frequently raised about the use of machine learning and artificial intelligence in clinical settings [17, 37–39]. This study found that incorporating predictors preceding an encounter by over a year provided only a small improvement in predictive performance. Thus, health systems could maintain strong overall risk identification without under-capturing variables for more recent enrollees by implementing a prediction model that excludes more distant predictors.

Variable importance analyses can provide a valuable tool for the development and implementation of prediction models in a variety of scenarios. For example, when transporting prediction models between settings, there may be concern that certain data elements are unavailable or incompletely captured in the new setting. If variables corresponding to such data elements are found to be important, investigators must carefully analyze the potential impacts on model performance.

Assessing importance of predictors that are expensive or intrusive to collect can provide guidance on which variables or variable groups to prioritize for collection. Variable importance analyses could also be used to investigate concerns about equity in clinical prediction models, e.g., by quantifying the predictive contribution of variables expected to be most affected by structural racism and other institutionalized systems of disadvantage. In all cases, the predictiveness measures used to quantify variable importance should be selected by the investigator based on the scientific question of interest. For example, in settings where risk predictions are used to target interventions, the relative costs of false positives versus false negatives may suggesting prioritizing PPV over sensitivity or vice versa.

There are several opportunities for future research to extend the work of this study. First, this analysis included data from integrated health systems with access to both clinical records and health insurance claims data. The importance of clinical predictors from different time periods may vary for health systems with limited access to external claims data. Second, we examined variable importance for 90-day self-harm outcomes. The predictive value of risk factors measured at different time periods preceding the prediction instance may vary for different event horizons. Examining importance for predicting outcomes over longer time horizons may inform the development of risk models used to target long-term interventions. Longer outcome time horizons may be particularly relevant in settings where individuals are assessed for self-harm risk at a single time point, as opposed to being assessed at multiple visits over time [40]. Third, while we did evaluate variable importance for predicting the risk of self-harm within subgroups defined by race and ethnicity, we were unable to perform a similar stratified analysis for suicide death due to the limited number of events in some subgroups. It may also be of interest to examine importance within patient subgroups defined by other variables such as type of insurance coverage. Prior to clinical use of a prediction model, it is imperative to assess performance in subgroups of interest to ensure implementation does not lead to inequitable allocation of health care resources [17]. Finally, while we focused on the importance of temporal predictors in this work, the analysis framework we employ here represents a general approach to evaluating variable importance, and analysis of other predictor categories may be of interest.

## 5. CONCLUSION

Self-harm risk prediction models often use predictors capturing up to five years of information on diagnoses, dispensed medications, and responses to the 9th item of the Patient Health Questionnaire. We found that the most recent three months of mental health–specific features were highly important for predicting the risk of non-fatal and fatal self-harm; removing these predictors resulted in a drop in AUC from 0.85 (all predictors) to 0.78. These findings suggest that rapid capture of recent data and integration into health records is crucial for predicting self-harm risk. We also demonstrated that the framework of algorithm-agnostic variable importance can be used to answer informatics questions with implications for the implementation of risk prediction models.

## AUTHOR CONTRIBUTIONS

**C.J.W.** Conceptualization, Formal analysis, Investigation, Methodology, Software, Validation, Visualization, Writing – original draft, Writing - review and editing. **B.D.W.** Conceptualization, Investigation, Methodology, Software, Supervision, Validation, Writing – original draft, Writing - review and editing. **S.M.S.** Conceptualization, Data curation, Writing - review and editing. **G.E.S.** Conceptualization, Data curation, Writing - review and editing. **K.J.C.** Conceptualization, Data curation, Writing - review and editing. **R.Y.** Conceptualization, Writing - review and editing. **B.K.A.** Conceptualization, Data curation, Writing - review and editing. **Y.D.** Conceptualization, Data curation, Writing - review and editing. **F.L.L.** Conceptualization, Data curation, Writing - review and editing. **R.C.R.** Conceptualization, Data curation, Writing - review and editing. **R.A.Z.** Conceptualization, Data curation, Writing - review and editing. **M.C.** Conceptualization, Writing - review and editing. **R.D.W.** Conceptualization, Writing - review and editing. **R.Y.C.** Conceptualization, Data curation, Formal analysis, Funding acquisition, Investigation, Methodology, Supervision, Validation, Writing – original draft, Writing - review and editing.

## DATA AVAILABILITY

The datasets generated and analyzed during this study are not publicly available because they contain detailed information from the electronic health records in the health systems participating in this study and are governed by HIPAA. Data are however available from the authors upon reasonable request, with permission of all health systems involved and fully executed data use agreement.

## Supplementary material

### SETTING THE *X*-AXIS SCALE FOR FIGURES (UNSTRATIFIED ANALYSIS)

For each combination of visit type and outcome, we calculated the maximum achievable variable importance using the performance of the full model including predictors from all time periods. For AUC, the maximum achievable importance is the difference between the AUC of the full model and 0.5 (the AUC of a null model). For sensitivity at a given cut-point, the maximum importance is the difference between the sensitivity of the full model and the proportion of visits flagged using that cut-point. For PPV, the maximum importance is the difference between the PPV of the full model and the overall event rate. In each figure, the upper limit of the *x*-axis is set to the maximum achievable importance.

### VARIABLE IMPORTANCE WITHIN SUBGROUPS

In addition to the primary analyses in the four outcome-setting pairs, we also performed analyses in subgroups defined by self-reported race and ethnicity. Due to the small number of suicide deaths in some subgroups, we performed stratified analyses only for self-harm, in both mental health and general medical settings. Race/ethnicity categories included White, Hispanic, Black/African American, Asian, American Indian/Alaskan Native, Native Hawaiian/Pacific Islander, multiracial or other race, and no race or ethnicity recorded. For each outcome-setting pair, penalized regression models were fit using all visits. Subgroup-specific estimates of predictiveness were then computed using only visits corresponding to that subgroup. For sensitivity and specificity, subgroup-specific risk score quantiles were used.

Performance of risk prediction models using predictors from all time periods is reported in Tables S3 and S4 Variable importance estimates are shown in Figures S5–S8.

### SUPPLEMENTARY TABLES AND FIGURES

**Table S1:**
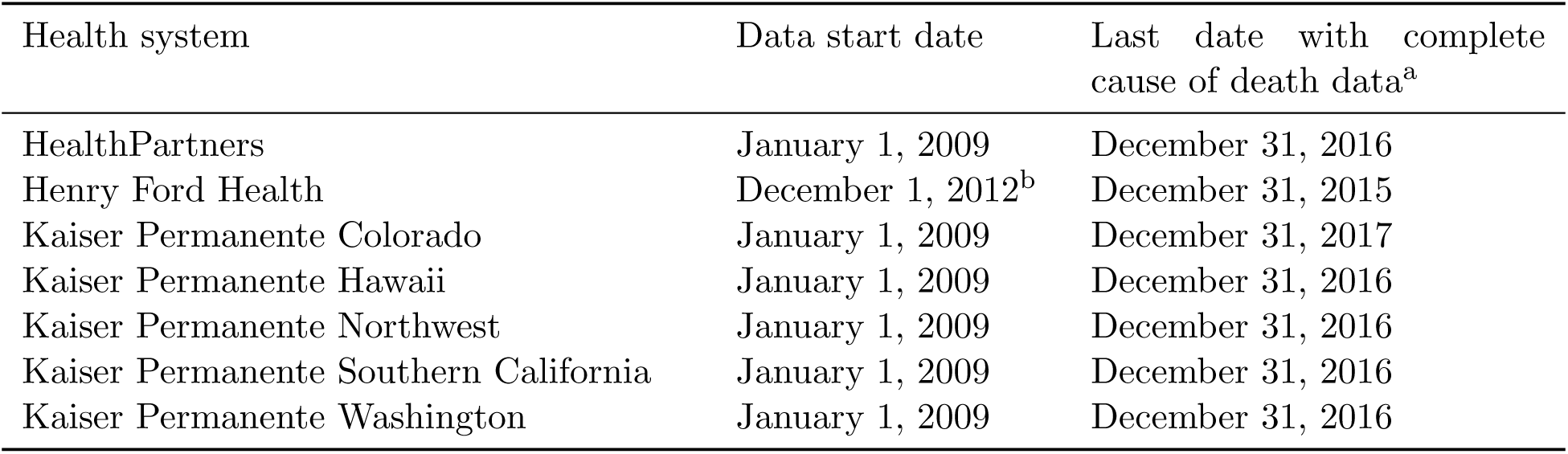
Data availability dates for participating sites. ^a^The study sample includes visits up to September 30 of the year with complete capture of cause of death data to allow for 90 days follow-up after mental health visits. For example, visits through September 30, 2016 are included for health systems with cause of death data complete through December 31, 2016. ^b^Only visits that occurred after the implementation of a new electronic health records system at Henry Ford were included in the sample.

| Health system | Data start date | Last date with complete cause of death data<sup>a</sup> |
| --- | --- | --- |
| HealthPartners | January 1, 2009 | December 31, 2016 |
| Henry Ford Health | December 1, 2012<sup>b</sup> | December 31, 2015 |
| Kaiser Permanente Colorado | January 1, 2009 | December 31, 2017 |
| Kaiser Permanente Hawaii | January 1, 2009 | December 31, 2016 |
| Kaiser Permanente Northwest | January 1, 2009 | December 31, 2016 |
| Kaiser Permanente Southern California | January 1, 2009 | December 31, 2016 |
| Kaiser Permanente Washington | January 1, 2009 | December 31, 2016 |

**Figure S1:**
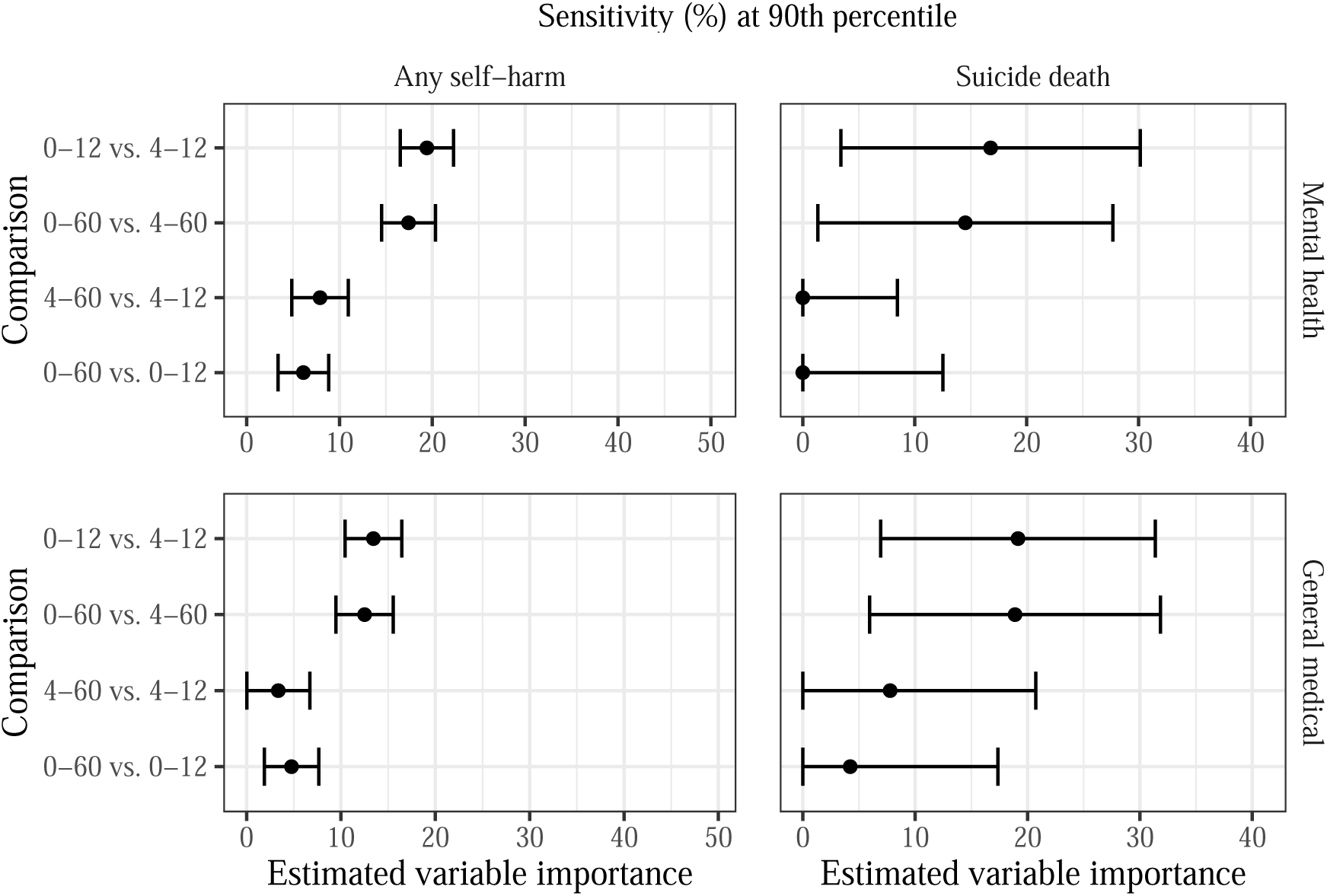
Estimated variable importance for temporal predictor groups in terms of **sensitivity at the 90th percentile** of risk scores. Note the different *x*-axis scales for each outcome-setting pair, which are based on the estimated maximum possible variable importance (see Supplementary Material for details).

**Table S2:** Overview of variables included in prediction models. MH: mental health. ^a^Each temporal predictor is a binary indicator of presence/absence in 0–3 months, 3 months – 1 year, or 1–5 years prior to the index visit.

| Base predictors |  |
| --- | --- |
| Age | Sex |
| Race | Hispanic ethnicity |
| Medicaid coverage at visit | Commercial insurance coverage at visit |
| Private pay insurance at visit | State-subsidized insurance at visit |
| Self-funded insurance at visit | Medicare insurance at visit |
| Other insurance at visit | High-deductible insurance at visit |
| Median household income <\$25k at visit<br>(census-based) | Median household income <\$40k at visit<br>(census-based) |
| Neighborhood <25% college educated at<br>visit (census-based) |  |
| Temporal predictors<sup>a</sup> |  |
| Depression diagnosis | Anxiety diagnosis |
| Bipolar diagnosis | Schizophrenia diagnosis |
| Other psychological disorder diagnosis | Dementia diagnosis |
| ADD diagnosis | ASD diagnosis |
| Personality disorder diagnosis | Alcohol use disorder diagnosis |
| Drug use disorder diagnosis | PTSD diagnosis |
| Eating disorder diagnosis | Traumatic brain injury diagnosis |
| Antidepressant prescription fill | Benzodiazepine prescription fill |
| Hypnotic prescription fill | Second generation antipsychotic<br>prescription fill |
| Inpatient encounter with MH diagnosis | Outpatient MH specialty visit |
| Emergency/urgent care encounter<br>MH diagnosis | Any self-inflicted injury/poisoning |
| Self-inflicted lacerative violent injury | Other self-inflicted violent injury |
| Any injury/poisoning diagnosis | Natal delivery diagnosis |
| Modal PHQ9 9th item response | Maximum PHQ9 9th item response |
| Number of PHQ9 9th item responses |  |

**Table S3:** Performance of any self-harm (fatal and non-fatal) prediction models including predictors from all time periods in the mental health setting.

|  | AUC | Sensitivity (%) |  |  |
| --- | --- | --- | --- | --- |
|  |  | 90<sup>th</sup> perc. | 95<sup>th</sup> perc. | 99<sup>th</sup> perc. |
| American Indian/Alaska Native | 0.846 | 58.1 | 41.6 | 11.6 |
| Asian | 0.840 | 56.6 | 42.9 | 16.3 |
| Black/African American | 0.827 | 53.9 | 40.6 | 16.4 |
| Native Hawaiian/Pacific Islander | 0.802 | 49.6 | 37.3 | 16.7 |
| White | 0.854 | 61.1 | 46.1 | 19.2 |
| Multiple or other races | 0.849 | 55.9 | 45.0 | 20.1 |
| Hispanic | 0.851 | 58.7 | 44.0 | 18.8 |
| No race/ethnicity indicated | 0.807 | 55.0 | 42.4 | 18.0 |

**Figure S2:**
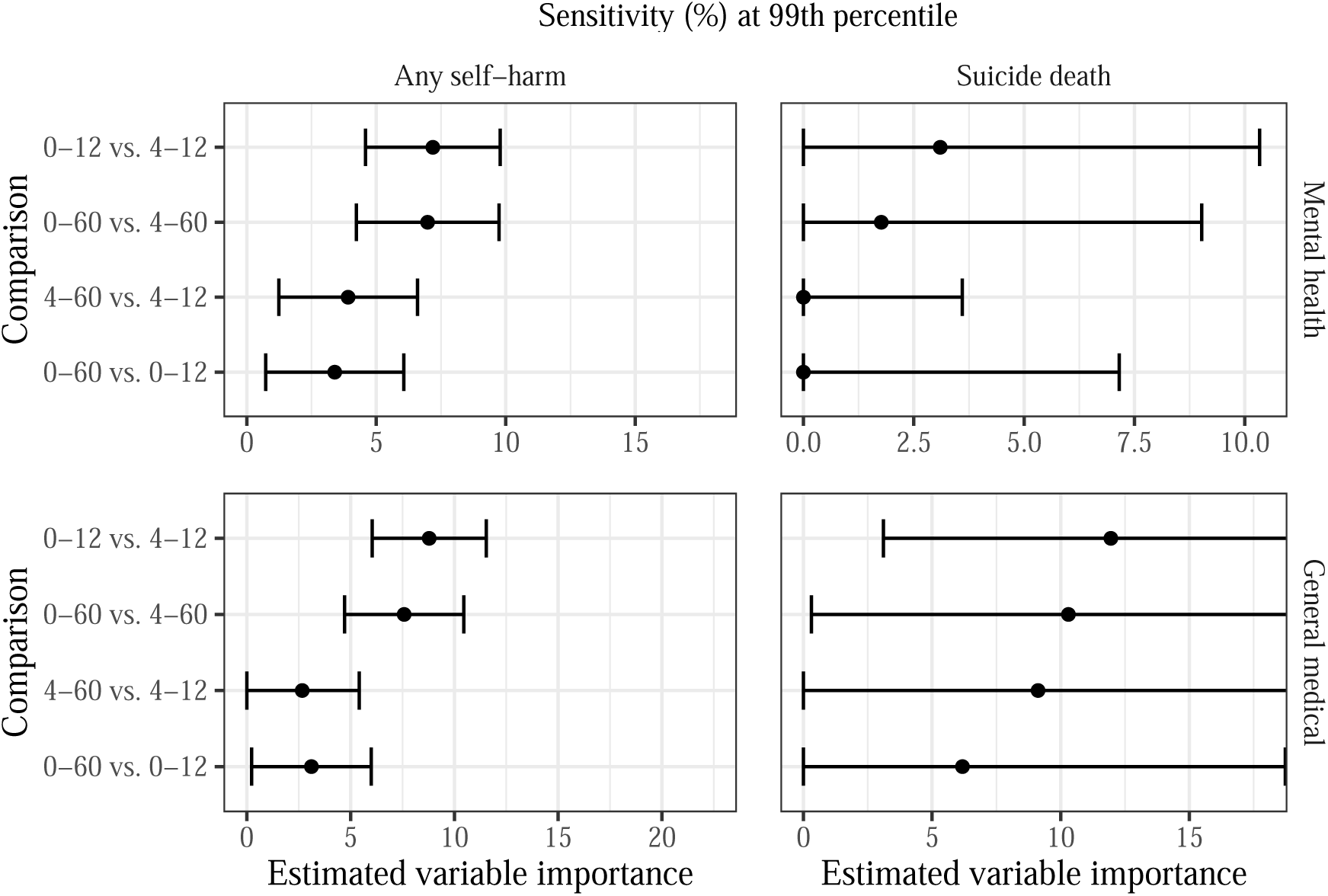
Estimated variable importance for temporal predictor groups in terms of **sensitivity at the 99th percentile** of risk scores. Note the different *x*-axis scales for each outcome-setting pair, which are based on the estimated maximum possible variable importance (see Supplementary Material for details).

**Table S4:** Performance of any self-harm (fatal and non-fatal) prediction models including predictors from all time periods in the general medical setting.

|  | AUC | Sensitivity (%) |  |  |
| --- | --- | --- | --- | --- |
|  |  | 90<sup>th</sup> perc. | 95<sup>th</sup> perc. | 99<sup>th</sup> perc. |
| American Indian/Alaska Native | 0.845 | 61.1 | 49.4 | 14.1 |
| Asian | 0.808 | 55.6 | 44.8 | 22.5 |
| Black/African American | 0.831 | 58.4 | 42.9 | 20.8 |
| Native Hawaiian/Pacific Islander | 0.820 | 54.5 | 34.5 | 17.9 |
| White | 0.829 | 59.5 | 47.3 | 24.2 |
| Multiple or other races | 0.798 | 50.9 | 42.2 | 28.1 |
| Hispanic | 0.839 | 60.8 | 48.2 | 23.2 |
| No race/ethnicity indicated | 0.810 | 58.2 | 47.3 | 20.4 |

**Figure S3:**
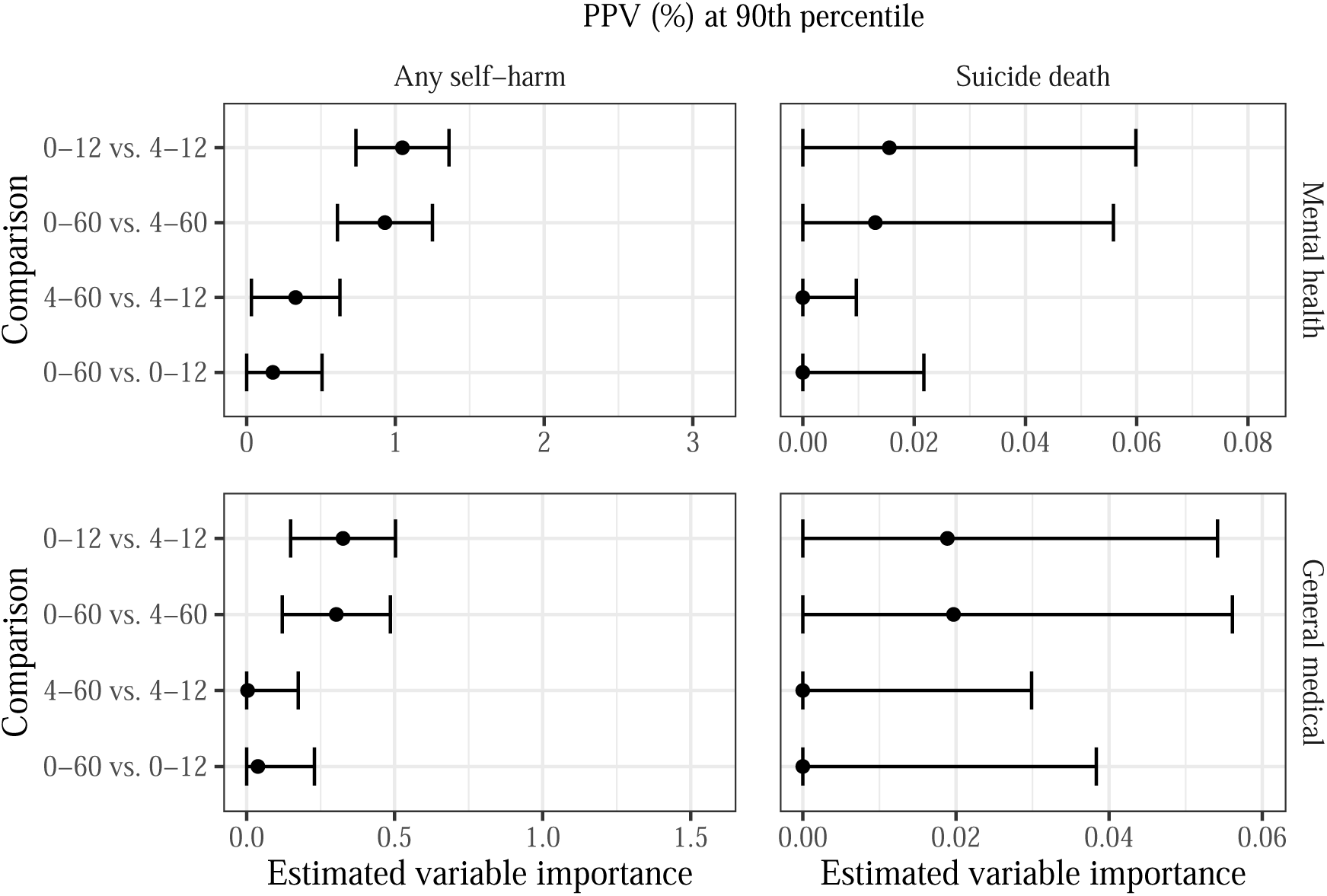
Estimated variable importance for temporal predictor groups in terms of **PPV at the 90th percentile** of risk scores. Note the different *x*-axis scales for each outcome-setting pair, which are based on the estimated maximum possible variable importance (see Supplementary Material for details).

**Figure S4:**
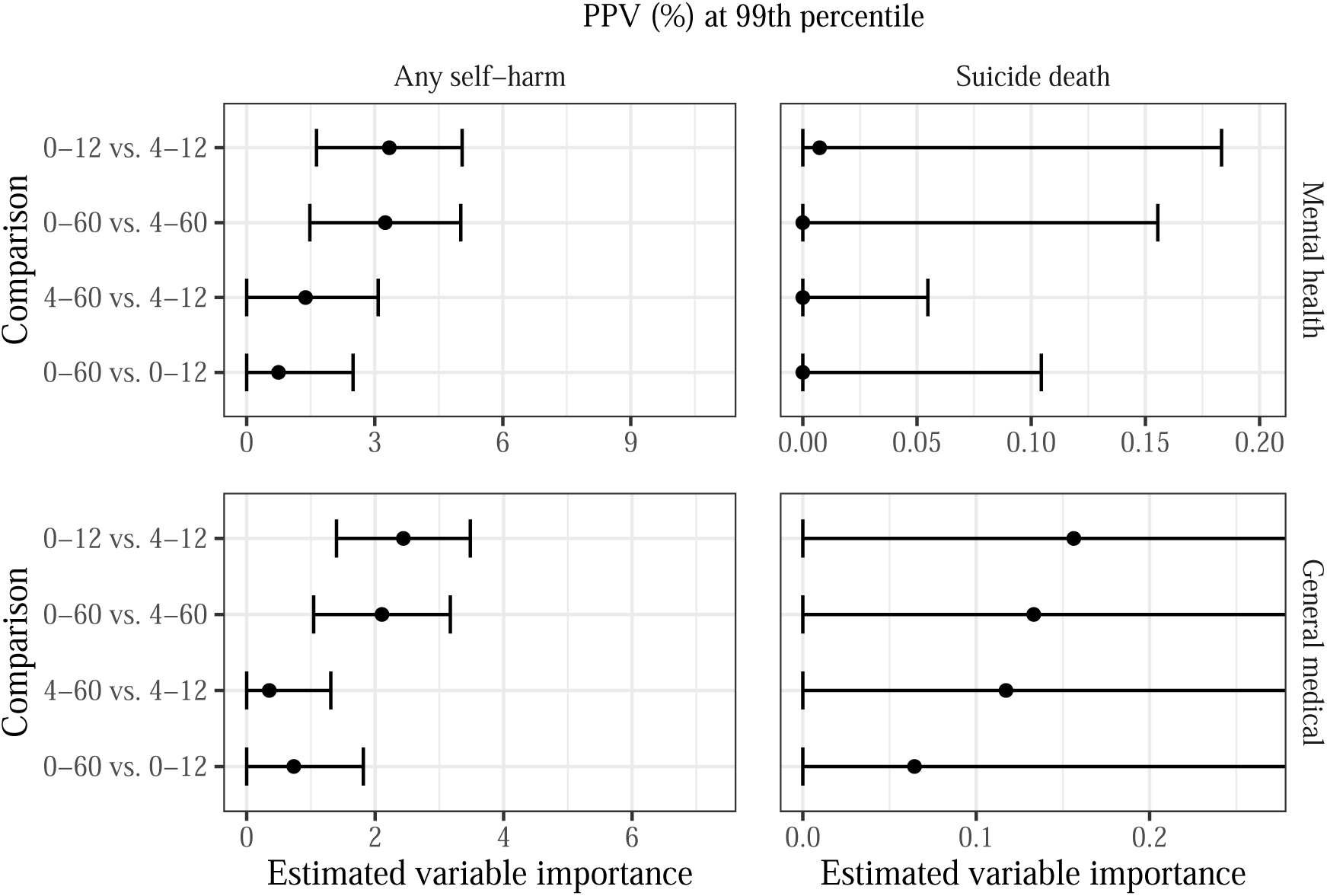
Estimated variable importance for temporal predictor groups in terms of **PPV at the 99th percentile** of risk scores. Note the different *x*-axis scales for each outcome-setting pair, which are based on the estimated maximum possible variable importance (see Supplementary Material for details).

**Figure S5:**
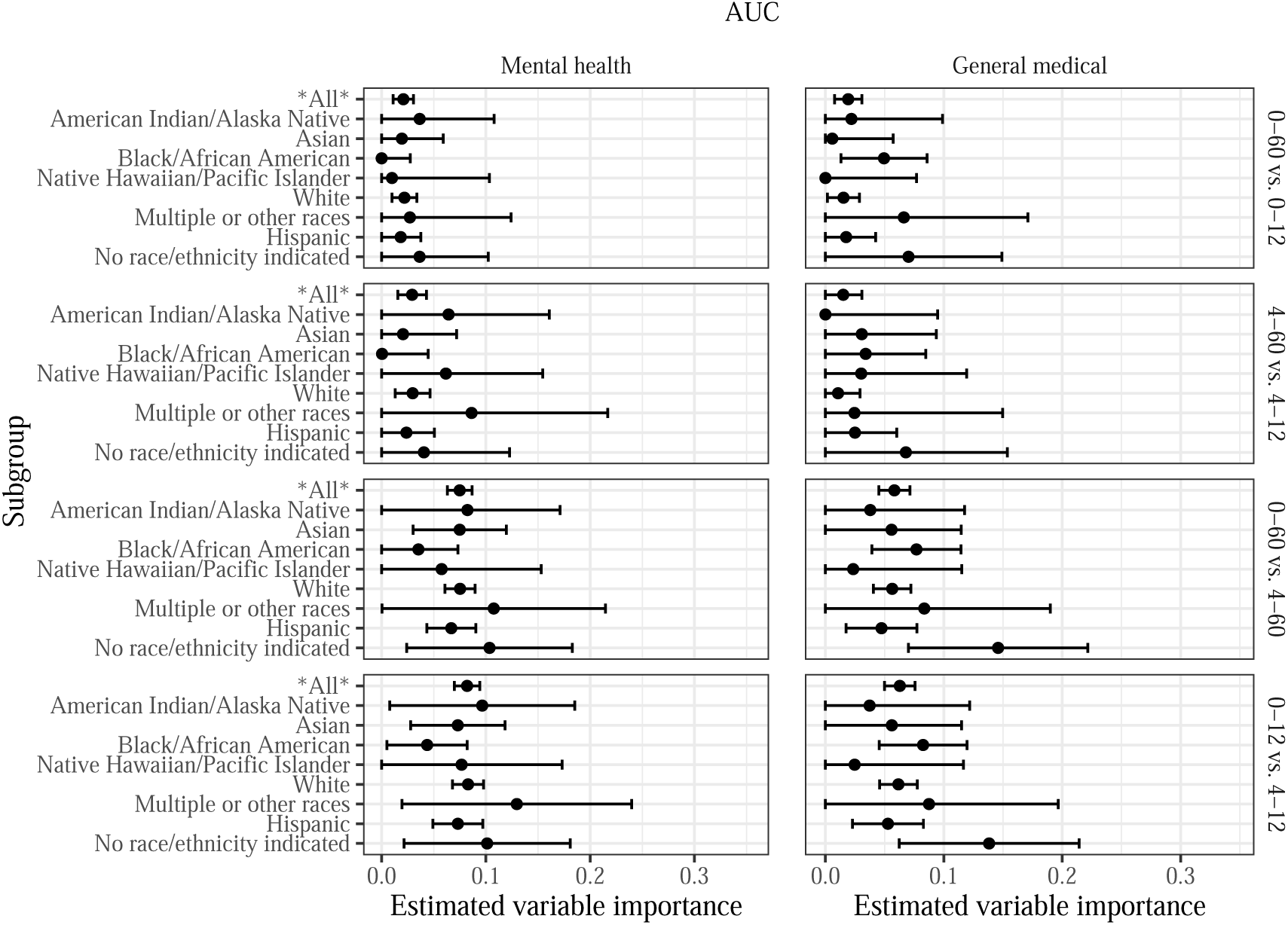
Estimated variable importance for temporal predictor groups in terms of **AUC**, stratified by race and ethnicity.

**Figure S6:**
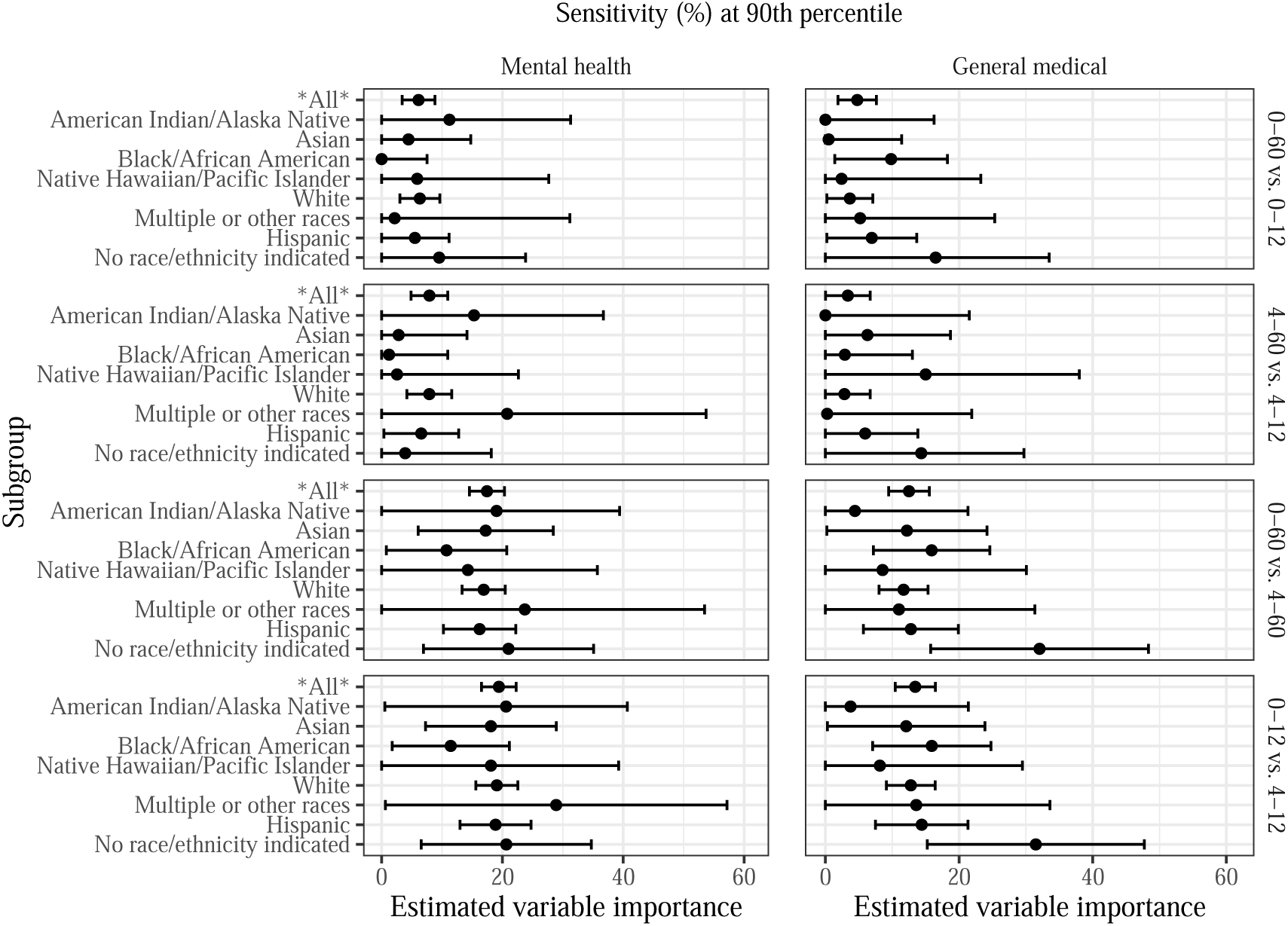
Estimated variable importance for temporal predictor groups in terms of **sensitivity at the 90th percentile** of risk scores, stratified by race and ethnicity.

**Figure S7:**
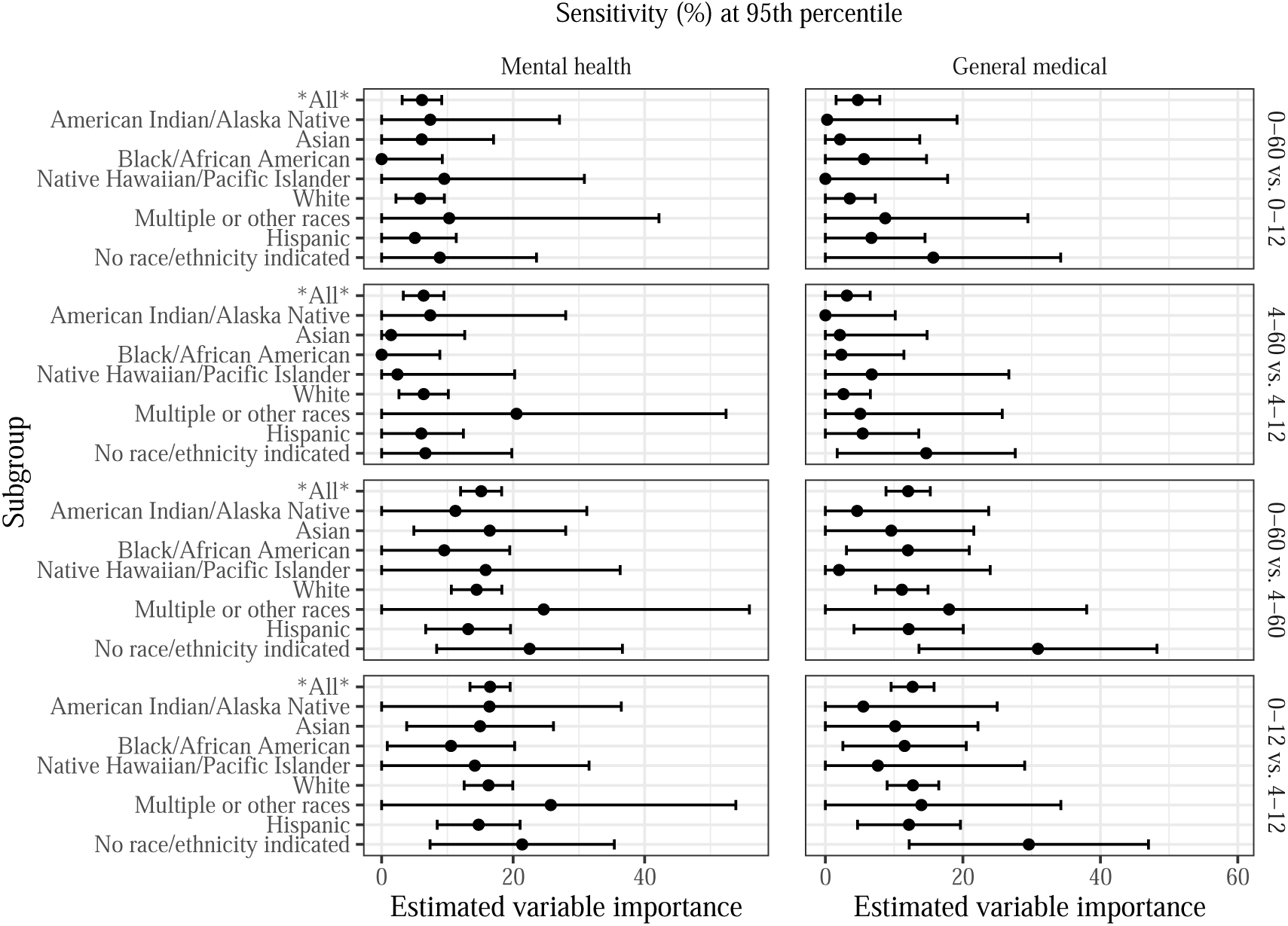
Estimated variable importance for temporal predictor groups in terms of **sensitivity at the 95th percentile** of risk scores, stratified by race and ethnicity.

**Figure S8:**
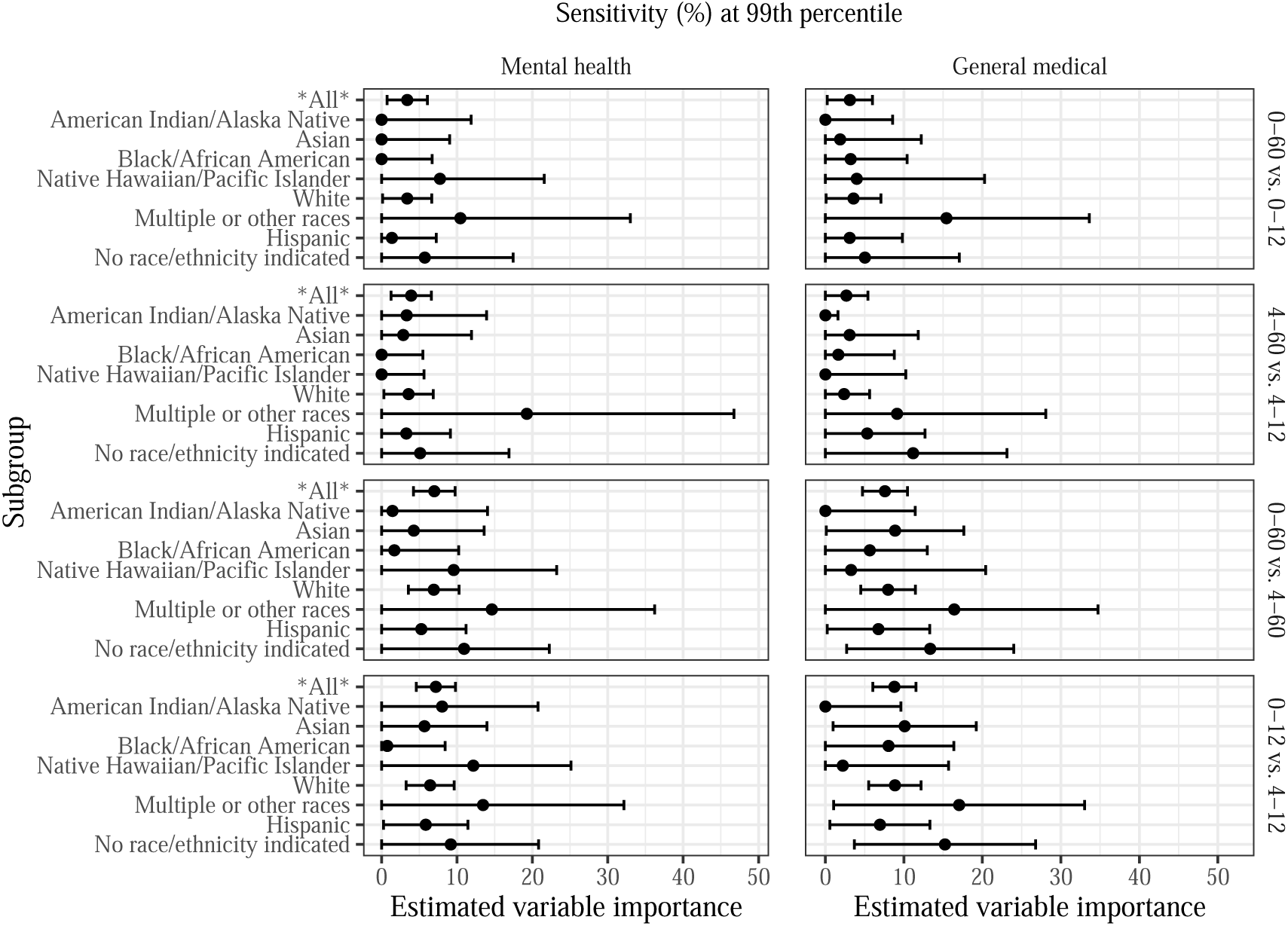
Estimated variable importance for temporal predictor groups in terms of **sensitivity at the 99th percentile** of risk scores, stratified by race and ethnicity.

